# Geldof Expert Consensus Classification of Perianal Fistulizing Crohn’s Disease: A Real-World Application in a Serial Fistula MRI Cohort

**DOI:** 10.1101/2024.02.03.24302160

**Authors:** Matthew Schroeder, Suha Abushamma, Alvin T. George, Ravella Balakrishna, John Hickman, Anusha Elumalai, Paul Wise, Maria Zulfiqar, Daniel R Ludwig, Anup Shetty, Satish E. Viswanath, Chongliang Luo, Shaji Sebastian, David H. Ballard, Parakkal Deepak

## Abstract

**Background and Aims:** Perianal fistulizing Crohn’s disease (CD-PAF) is an aggressive phenotype of Crohn’s disease (CD) defined by frequent relapses and disabling symptoms. A novel consensus classification system was recently outlined by Geldof et al. that seeks to unify disease severity with patient-centered goals but has not yet been validated. We aimed to apply this to a real-world cohort and identify factors that predict transition between classes over time.

**Methods:** We identified all patients with CD-PAF and at least one baseline and one follow-up pelvic (pMRI). Geldof Classification, disease characteristics, and imaging indices were collected retrospectively at time periods corresponding with respective MRIs.

**Results:** We identified 100 patients with CD-PAF of which 96 were assigned Geldof Classes 1 – 2c at baseline. Most patients (78.1%) started in Class 2b, but changes in classification were observed in 52.1% of all patients. Male sex (72.0%, 46.6%, 40.0%, p = 0.03) and prior perianal surgery (52.0% vs 44.6% vs 40.0%, p = 0.02) were more frequently observed in those with improved. Baseline pMRI indices were not associated with changes in classification, however, greater improvements in mVAI, MODIFI-CD, and PEMPAC were seen among those who improved. Linear mixed effect modeling identified only male sex (−0.31, 95% CI −0.60 to −0.02) with improvement in class.

**Conclusion:** Geldof classification highlights the dynamic nature of CD-PAF over time, however, our ability to predict transitions between classes remains limited and requires prospective assessment. Improvement in MRI index scores over time was associated with a transition to lower Geldof classification.

## Introduction

Crohn’s disease (CD) is characterized by transmural inflammation of the gastrointestinal (GI) mucosa^1^. Perianal-fistulizing CD (CD-PAF) is a distinct, aggressive phenotype that occurs in approximately 20-30% of patients^2, 3^ and is independently associated with poor quality of life, frequent hospitalizations, and need for multiple surgical interventions such as fecal diversion and total proctectomy^4, 5^. Some fistulas are refractory to both medical and surgical management. Clinical targets are evolving and include total symptom burden, total number of draining fistulas, closure of external orifices, and radiographic transition from patent fistulous tracts to a fibrotic appearance^6–8^.

Traditional anatomic descriptors such as Park’s classification describe the relation of the fistulous tract to the external and internal anal sphincters but fail to provide detail regarding complicating features^9^. The American Gastroenterological Association (AGA) classification provides a pragmatic division of fistula into simple and complex morphologies based on tract anatomy (low vs high), external openings (single vs multiple), and the presence of underlying proctitis, abscess, stricture, and ano-vaginal extension^10^. While complex fistulas portend worse prognosis in some studies^11, 12^, the classification scheme alone lacks definitive clinical guidance. Pelvic MRI (pMRI) has emerged as the preferred modality for perianal fistula evaluation and is well suited to the delineation of specific anatomic features. CD-PAF pMRI scoring systems have been developed to quantify perianal disease severity and treatment response^13^.

Recently a new classification system has been developed through a modified nominal group technique expert consensus process, outlined by Geldof et al. in 2022^14^. This classification system generates a broad overarching framework unifying disease severity and patient-centered treatment goals using four distinct classes, extending beyond the emphasis on fistula anatomy, morphology, and activity of current clinical and pMRI based classifications and scoring systems. While this proposed classification has been suggested as a framework to standardize clinical practice and better organize patients with CD-PAF for clinical trials, it has yet to be validated in a real-world cohort.

We aimed to evaluate the Geldof classification system using a real-world cohort of patients with CD-PAF who underwent serial pMRI and identify factors that may predict transition between classes over time.

## Methods

Consecutive CD-PAF patients having previously undergone at least two pMRI with fistula protocol at our quaternary referral center from November 2^nd^, 2011, to May 5^th^, 2022, were reviewed for inclusion in the study using a pre-existing imaging cohort. The retrospective imaging cohort included adult and pediatric patients with radiographic evidence of perianal fistula based on pMRI and an underlying diagnosis of CD as determined by the primary inflammatory bowel disease (IBD)-focused Gastroenterologist. Patients without adequate pMRI protocol to evaluate for perianal fistulizing disease and those without associated clinician follow-up at our center were excluded. The institutional review board of Washington University School of Medicine in St. Louis approved the study.

Baseline demographic and CD-specific data including anatomic, radiographic, and symptom-based classifications of perianal disease, prior medical and surgical management, and disease-related complications were collected retrospectively at time of patients’ first pMRI. Baseline was defined as time of patient’s first pMRI. First and second follow-up intervals were defined by dates correlating with each subsequent pMRI and corresponding clinical data was collected within a three-month period before or after imaging. Active medication use was defined as patients’ medication regimen following initial pMRI to reflect reactive changes following new diagnosis of perianal disease. A minimum of two pMRI were required in the study design to observe changes in Geldof classification and MRI indices. Patients with suspected CD-PAF underwent a baseline pMRI while those with known CD-PAF underwent serial pMRI based on the varying practice of the treating gastroenterologist or colorectal surgeon.

Geldof classifications **(Figure 1)** were assigned at baseline and at each follow-up interval. An initial protocol was developed using guidance published by Geldof et al. as part of the defined consensus criteria^14^. The full study definition used to define each study patient at each time point for the Geldof classification, is detailed in **Appendix A**. A sample of 20 patients from our imaging cohort were independently reviewed by a Gastroenterology fellow and an IBD-focused Gastroenterologist. Geldof classifications were assigned at each time. Discrepancies were reviewed and settled by a second IBD-focused Gastroenterologist, and the protocol was revised. Further assignments were made by the Gastroenterology fellow. While Geldof classifications are associated with specific procedural interventions, execution of procedures was not required for classification assignment.

**Figure 1:**
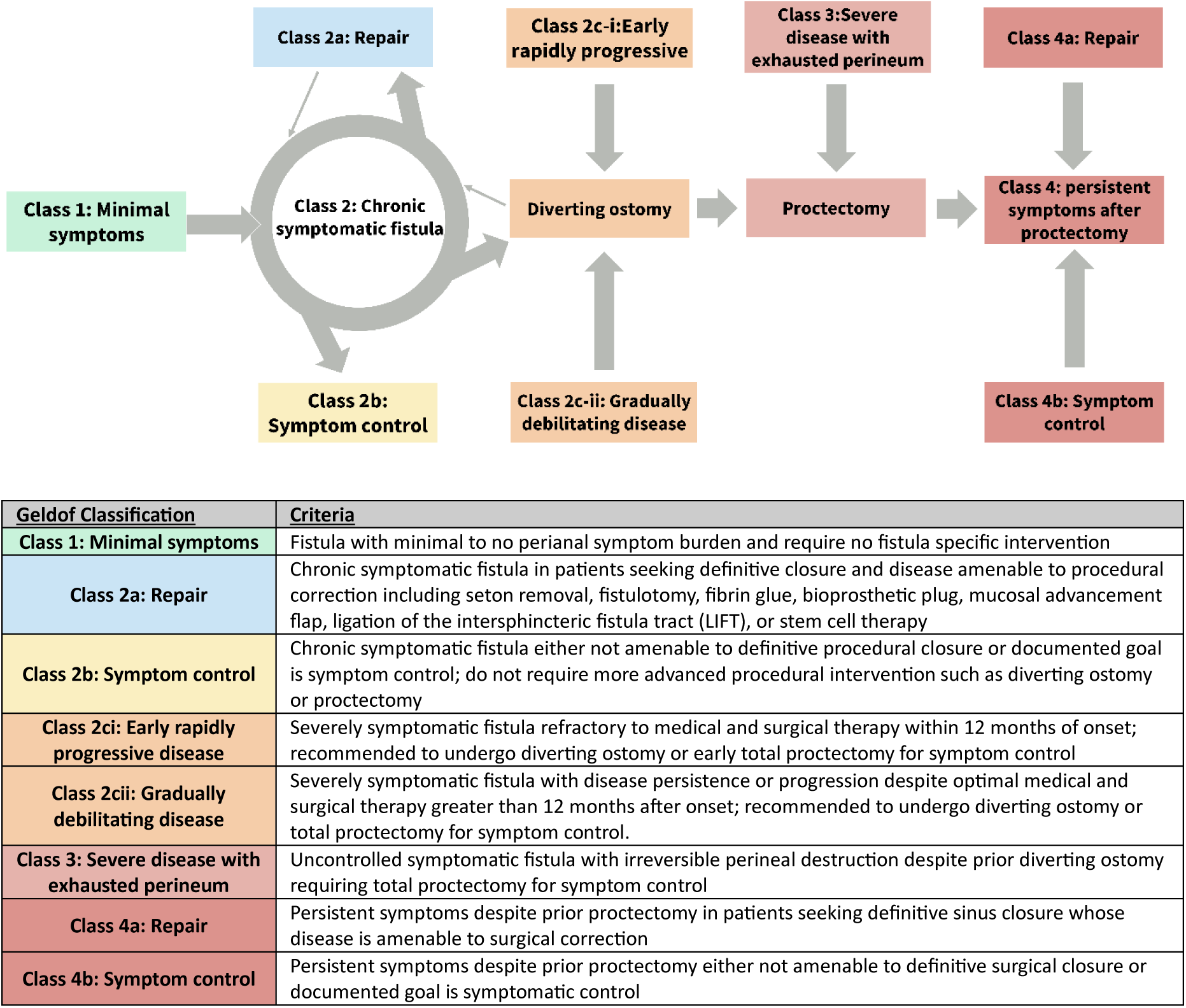
Geldof expert consensus classification scheme

Each pMRI was reviewed by one of four independent fellowship trained abdominal radiologists who regularly interpret pMRI with fistula protocol in CD-PAF patients as part of their clinical practice. For retrospective imaging review preparation, the study radiologists convened for a focused 1.5-hour training session. During this session, they reviewed the definitions of the MAGNIFI-CD, mVAI, PEMPAC pMRI scoring systems, and training sets of CD-PAF cases not included in the patient imaging cohort^15–17^.

Radiologists were blinded to associated clinical information except for age and sex. The radiologists had access to both the baseline and follow-up pMRI and were instructed to complete the baseline before interpreting each sequential follow-up. The study radiologists evaluated individual subcomponents for MRI indices (MAGNIFI-CD, mVAI, PEMPAC), towards calculated total weighted scores (**Appendix B**)^15–17^. Radiologist performance and reliability in scoring the pMRI in this dataset has been reported^18^. Changes in MRI scores were calculated by subtracting the most recent score from the preceding score.

In preliminary statistical analysis, patients were stratified by an initial change in Geldof classification between baseline and the first follow-up as worse (transition to a higher stage), stable, or improved (downgrading to a lower stage). Patients with Geldof Class 3 or 4 at baseline were excluded from further analysis as they are unable to return to lower classes in subsequent follow-ups. Continuous and categorical patient characteristics at baseline were compared between the three groups. Clinical outcomes outside of change in Gelof classification were not included in analysis as expected clinical course including recommended surgical management are incorporated into initial assignment. The associations between baseline patient characteristics and Geldof change were tested by univariate analysis of variance (ANOVA). Given the hierarchical and dynamic nature of the classification, Geldof class changes were handled as continuous variables (i.e., Geldof levels: 1 < 2a < 2b < 2c-i = 2c-ii < 3 < 4a = 4b, if change in class 2a to 2c-ii, then change = +2). The cohort data is presented as mean for normally distributed data and median for non-Gaussian distributed data. Variables with a p < 0.05 by ANOVA were incorporated into a multivariate regression model for the longitudinal Geldof change from baseline to follow-up 1, or follow-up 1 to follow-up 2. A linear mixed-effect model with patient-level random intercepts was fitted. R software (R Core Team, 2022) and IBM SPSS software (IBM Corp, 2022) were used for statistical analysis.

## Results

We identified 100 patients with perianal fistulizing CD (CD-PAF) and at least two pMRI with fistula protocol during the study period (**Figure 2**). Each patient had at least 2 pMRI required for appropriate fistula evaluation. Fifty-eight (58%) patients had a third pMRI available for review, for a total of 258 MRIs in the 100 CD-PAF patients. The median time between baseline MRI and follow-up MRI 1 was 10.5 months (IQR 5.5 – 18.8 months). The median time between follow-up MRI 1 and follow-up MRI 2 was 12.3 months (IQR 6.0 – 25.5 months). At baseline, two patients each were assigned to Class 3 and Class 4 and hence excluded from further analysis as they are unable to return to lower classes in subsequent follow-ups. The remaining 96 patients (Class 1 – Class 2c) were included in analysis of variance (**Table 1**). Patients were 52.1% male, 72.9% White, median age 28.5 years (IQR 18.6 – 43.9 years), median time since CD diagnosis 4.9 years (0.3 – 15.1 years), and median time since onset of perianal fistulae 1.1 years (0 – 7.8 years) prior to initial pelvic MRI. At baseline, 43 (44.8%) patients first developed symptoms of perianal fistula in the 6 months prior to initial pRMR. There were 25 (26.0%) patients newly diagnosed with CD in the 6 months prior to inclusion in the study.

**Figure 2:**
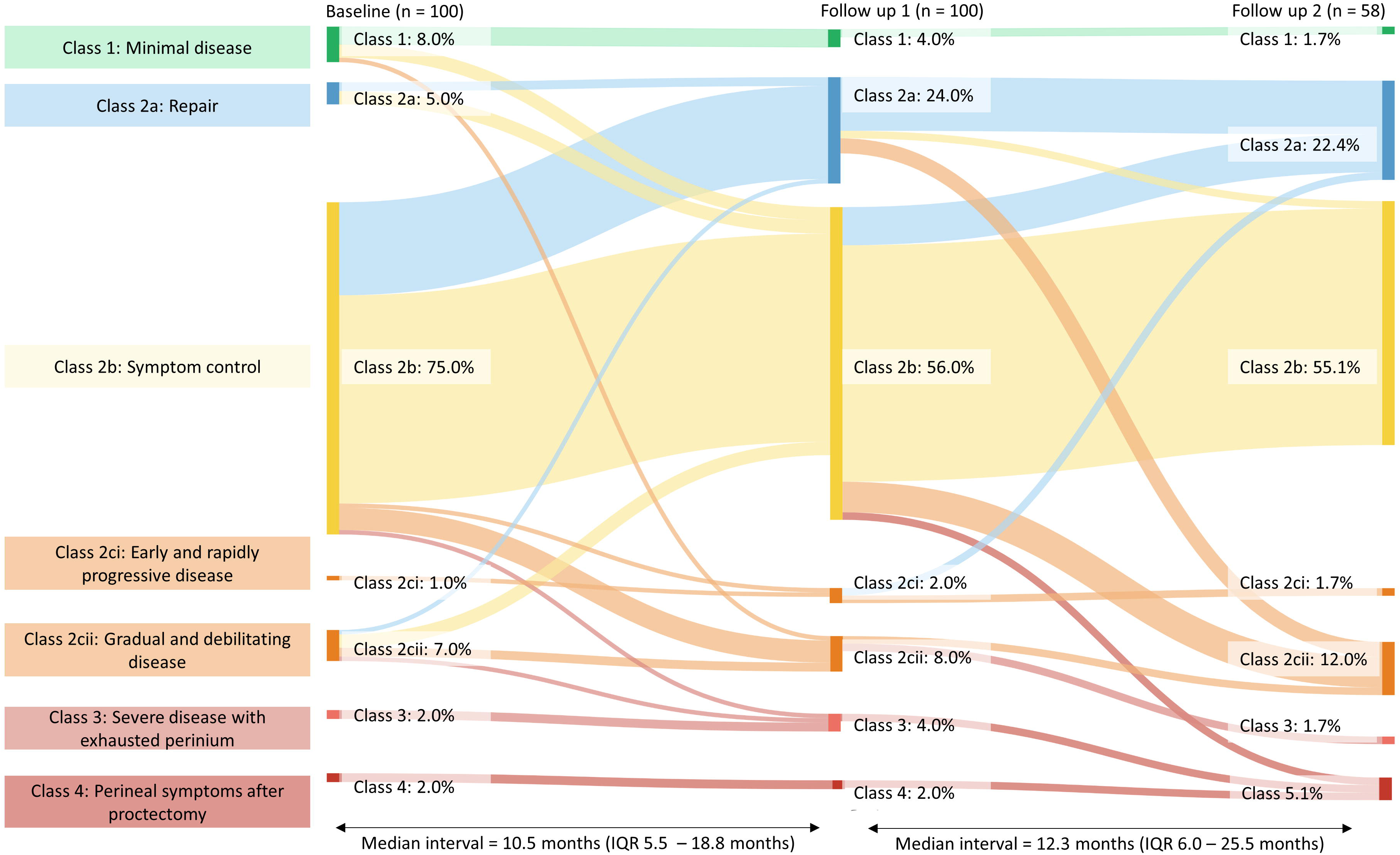
Changes in Geldof Classification between baseline and follow-up intervals

**Table 1:**
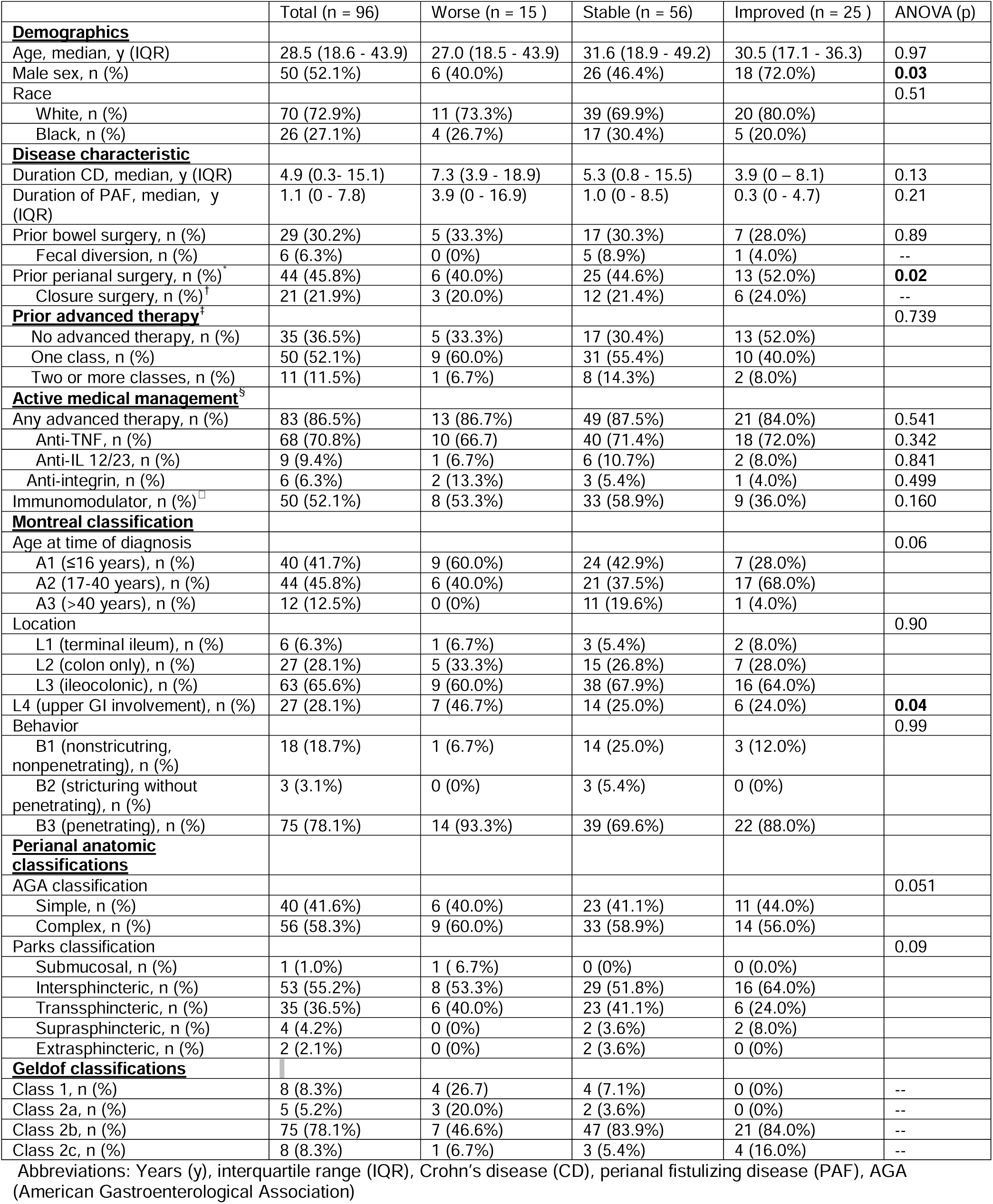
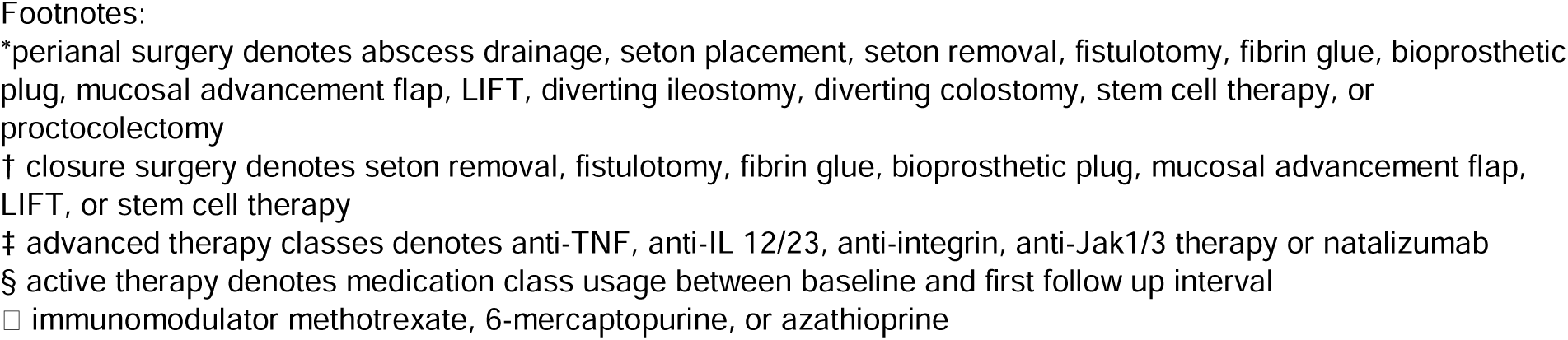
Baseline characteristics of cohort stratified by changes in Geldof classification between baseline and first follow-up pMRI.

|  | Total (n = 96) | Worse (n = 15 ) | Stable (n = 56) | Improved (n = 25 ) | ANOVA (p) |
| --- | --- | --- | --- | --- | --- |
| **Demographics** |  |  |  |  |  |
| Age, median, y (IQR) | 28.5 (18.6 - 43.9) | 27.0 (18.5 - 43.9) | 31.6 (18.9 - 49.2) | 30.5 (17.1 - 36.3) | 0.97 |
| Male sex, n (%) | 50 (52.1%) | 6 (40.0%) | 26 (46.4%) | 18 (72.0%) | **0.03** |
| Race |  |  |  |  | 0.51 |
| White, n (%) | 70 (72.9%) | 11 (73.3%) | 39 (69.9%) | 20 (80.0%) |  |
| Black, n (%) | 26 (27.1%) | 4 (26.7%) | 17 (30.4%) | 5 (20.0%) |  |
| **Disease characteristic** |  |  |  |  |  |
| Duration CD, median, y (IQR) | 4.9 (0.3- 15.1) | 7.3 (3.9 - 18.9) | 5.3 (0.8 - 15.5) | 3.9 (0 – 8.1) | 0.13 |
| Duration of PAF, median, y (IQR) | 1.1 (0 - 7.8) | 3.9 (0 - 16.9) | 1.0 (0 - 8.5) | 0.3 (0 - 4.7) | 0.21 |
| Prior bowel surgery, n (%) | 29 (30.2%) | 5 (33.3%) | 17 (30.3%) | 7 (28.0%) | 0.89 |
| Fecal diversion, n (%) | 6 (6.3%) | 0 (0%) | 5 (8.9%) | 1 (4.0%) | -- |
| Prior perianal surgery, n (%)<sup>†</sup> | 44 (45.8%) | 6 (40.0%) | 25 (44.6%) | 13 (52.0%) | **0.02** |
| Closure surgery, n (%)<sup>†</sup> | 21 (21.9%) | 3 (20.0%) | 12 (21.4%) | 6 (24.0%) | -- |
| **Prior advanced therapy<sup>‡</sup>** |  |  |  |  | 0.739 |
| No advanced therapy, n (%) | 35 (36.5%) | 5 (33.3%) | 17 (30.4%) | 13 (52.0%) |  |
| One class, n (%) | 50 (52.1%) | 9 (60.0%) | 31 (55.4%) | 10 (40.0%) |  |
| Two or more classes, n (%) | 11 (11.5%) | 1 (6.7%) | 8 (14.3%) | 2 (8.0%) |  |
| **Active medical management<sup>§</sup>** |  |  |  |  |  |
| Any advanced therapy, n (%) | 83 (86.5%) | 13 (86.7%) | 49 (87.5%) | 21 (84.0%) | 0.541 |
| Anti-TNF, n (%) | 68 (70.8%) | 10 (66.7) | 40 (71.4%) | 18 (72.0%) | 0.342 |
| Anti-IL 12/23, n (%) | 9 (9.4%) | 1 (6.7%) | 6 (10.7%) | 2 (8.0%) | 0.841 |
| Anti-integrin, n (%) | 6 (6.3%) | 2 (13.3%) | 3 (5.4%) | 1 (4.0%) | 0.499 |
| Immunomodulator, n (%)<sup>□</sup> | 50 (52.1%) | 8 (53.3%) | 33 (58.9%) | 9 (36.0%) | 0.160 |
| **Montreal classification** |  |  |  |  |  |
| Age at time of diagnosis |  |  |  |  | 0.06 |
| A1 (≤16 years), n (%) | 40 (41.7%) | 9 (60.0%) | 24 (42.9%) | 7 (28.0%) |  |
| A2 (17-40 years), n (%) | 44 (45.8%) | 6 (40.0%) | 21 (37.5%) | 17 (68.0%) |  |
| A3 (>40 years), n (%) | 12 (12.5%) | 0 (0%) | 11 (19.6%) | 1 (4.0%) |  |
| Location |  |  |  |  | 0.90 |
| L1 (terminal ileum), n (%) | 6 (6.3%) | 1 (6.7%) | 3 (5.4%) | 2 (8.0%) |  |
| L2 (colon only), n (%) | 27 (28.1%) | 5 (33.3%) | 15 (26.8%) | 7 (28.0%) |  |
| L3 (ileocolonic), n (%) | 63 (65.6%) | 9 (60.0%) | 38 (67.9%) | 16 (64.0%) |  |
| L4 (upper GI involvement), n (%) | 27 (28.1%) | 7 (46.7%) | 14 (25.0%) | 6 (24.0%) | **0.04** |
| Behavior |  |  |  |  | 0.99 |
| B1 (nonstricturing, nonpenetrating), n (%) | 18 (18.7%) | 1 (6.7%) | 14 (25.0%) | 3 (12.0%) |  |
| B2 (stricturing without penetrating), n (%) | 3 (3.1%) | 0 (0%) | 3 (5.4%) | 0 (0%) |  |
| B3 (penetrating), n (%) | 75 (78.1%) | 14 (93.3%) | 39 (69.6%) | 22 (88.0%) |  |
| **Perianal anatomic classifications** |  |  |  |  |  |
| AGA classification |  |  |  |  | 0.051 |
| Simple, n (%) | 40 (41.6%) | 6 (40.0%) | 23 (41.1%) | 11 (44.0%) |  |
| Complex, n (%) | 56 (58.3%) | 9 (60.0%) | 33 (58.9%) | 14 (56.0%) |  |
| Parks classification |  |  |  |  | 0.09 |
| Submucosal, n (%) | 1 (1.0%) | 1 ( 6.7%) | 0 (0%) | 0 (0.0%) |  |
| Intersphincteric, n (%) | 53 (55.2%) | 8 (53.3%) | 29 (51.8%) | 16 (64.0%) |  |
| Transsphincteric, n (%) | 35 (36.5%) | 6 (40.0%) | 23 (41.1%) | 6 (24.0%) |  |
| Suprasphincteric, n (%) | 4 (4.2%) | 0 (0%) | 2 (3.6%) | 2 (8.0%) |  |
| Extrasphincteric, n (%) | 2 (2.1%) | 0 (0%) | 2 (3.6%) | 0 (0%) |  |
| **Geldof classifications** |  |  |  |  |  |
| Class 1, n (%) | 8 (8.3%) | 4 (26.7) | 4 (7.1%) | 0 (0%) | -- |
| Class 2a, n (%) | 5 (5.2%) | 3 (20.0%) | 2 (3.6%) | 0 (0%) | -- |
| Class 2b, n (%) | 75 (78.1%) | 7 (46.6%) | 47 (83.9%) | 21 (84.0%) | -- |
| Class 2c, n (%) | 8 (8.3%) | 1 (6.7%) | 3 (5.4%) | 4 (16.0%) | -- |

Abbreviations: Years (y), interquartile range (IQR), Crohn's disease (CD), perianal fistulizing disease (PAF), AGA (American Gastroenterological Association)
\*perianal surgery denotes abscess drainage, seton placement, seton removal, fistulotomy, fibrin glue, bioprosthetic plug, mucosal advancement flap, LIFT, diverting ileostomy, diverting colostomy, stem cell therapy, or proctocolectomy
† closure surgery denotes seton removal, fistulotomy, fibrin glue, bioprosthetic plug, mucosal advancement flap, LIFT, or stem cell therapy
‡ advanced therapy classes denotes anti-TNF, anti-IL 12/23, anti-integrin, anti-Jak1/3 therapy or natalizumab
§ active therapy denotes medication class usage between baseline and first follow up interval
□ immunomodulator methotrexate, 6-mercaptopurine, or azathioprine

Prior to the study period, most patients (63.5%) had received at least one class of biologic or small molecule therapy. Following initial MRI, 86.5% of patients were being managed with advanced therapies with a smaller fraction (52.1%) receiving concomitant immunomodulators. Before the first pMRI 45.8% of patients had undergone prior perianal related surgery, but only 21.9% had prior surgery directed towards fistula closure. Fistulas were most commonly complex by AGA criteria (58.3%) with intersphincteric (55.2%) and transsphincteric (36.5%) being the most observed Park’s classifications.

Baseline (**Table 1**), the large majority (78.1%; 75/96) of patients were classified as Geldof Class 2b (symptom control) with only 5.2% (5/96) suitable for repair (Class 2a) and 8.3% (8/96) classified as Class 2c. Minimal symptoms (Class 1) were reported in 8.3% (8/96). Over median study duration of 1.8 years (IQR 1.1 – 3.1 years), approximately half of patients (52.1%; 50/96) changed Geldof classifications at least once (**Figure 2**). Of the 83 patients at baseline with chronic symptomatic fistula not amenable to fistula closure (Class 2b - c), 27 (32.5%) transitioned to class 2a at some point during the study. Transition from symptom control (2b) to repair (2a) was the most frequently observed transition. When assigned class 2b at baseline or follow-up 1, 19.8% (26/131 instances) transited to Class 2a. Of the 27 patients classified as class 2a (at baseline or follow-up 1), ultimately 21/27 (77.8 %) underwent procedural intervention directed at definitive closure of fistula, with removal of non-cutting seton being the most common (57%; 12/21). Half (4/8) of patients with minimal or no symptoms (Class 1) later progressed to a symptomatic class. At baseline, 8.3% (8/96) of patients were classified as Class 2c with severe disease recommended for diverting ostomy or proctectomy. Ultimately, the proportion of patients with severe disease classified between Class 2c – Class 4 increased to 24.0% (23/96) during the study period. There were 18 patients who underwent diverting ostomy, , two who underwent proctectomy, and three who declined either diversion or proctectomy. Of the 18 patients who underwent ileostomy, four ( later underwent ileostomy take down and re-classified at class 2a on subsequent follow up.

Between baseline and the first follow-up interval, 15 patients transitioned to a worse Geldof classification, while 56 remained unchanged, and 25 improved. There was a significantly higher proportion of males in the improved group (72.0%, 18/25, p = 0.03) when compared to those who were stable (46.4%, 26/56) and worsened (40.0%, 6/15) (**Table 1**).

Prior history of perianal surgery was more frequently observed in the improved group compared to stable and worsened groups (52.0% vs 44.6% vs 40.0%, p = 0.02). Upper gastrointestinal involvement was more commonly seen in those with worsening Geldof classifications (46.7%, p = 0.04) when compared to those with stable (25.0%) and improved (24.0%) classifications. Complex fistulas trended towards worsening Geldof classification but did not achieve statistical significance (p = 0.051). There was no significant difference in prior medication class exposure, active medication use, age at diagnosis, age at enrollment, or fistula anatomy by Parks classification.

Initial radiographic features of perianal fistulas are presented in **Table 2**. Composite scores for each MRI index at baseline were calculated with median MAGNIFI-CD 14 (IQR 10 - 18) (25 possible total MAGNIFI-CD points), median mVAI 8.3 (IQR 4.7 – 12.2) (19.5 possible total mVAI points), and median PEMPAC 26 (IQR 15 – 31) (41 possible total PEMPAC points). The averaged MRI score was defined as MAGNIFI-CD/25 + mVAI/19.5 + PEMPAC/41. There was a small average improvement in each of the MRI indices between baseline and follow-up one, with mean change of MAGNIFI-CD −0.7 (σ 6.4), mVAI −1.2 (σ 4.8), PEMPAC −1.5 (σ 8.9), and averaged MRI score −0.0473 (0.220) (**Table 2**). There was no significant difference in baseline MRI index scores between those who improved, remained stable, or worsened at first follow up (**Table 2**). However, there was a greater reduction across all three scores: total MAGNIFI-CD (−2.3 vs −0.4 vs 1.0, p = 0.04), mVAI (−2.4 vs −0.9 vs −0.4, p = 0.03), PEMPAC scores (−4.1 vs −0.8 vs 0.0, p = 0.03), and averaged MRI score (−0.108 vs −0.0340 vs −0.00354) when comparing patients who improved vs. those who either remained stable or transitioned to a worse Geldof classification. On subcomponent analysis, only tract hyperintensity on T2 was significantly different between the three groups (p = 0.04). There was a trend towards higher frequency of ano-vaginal tract extension in patients with worsening classification (20.0%, p = 0.051) compared to those with stable (16.1%) and improved (4.0%) classification.

**Table 2:** Pelvic MRI characteristics stratified by changes in Geldof classification between baseline and first follow-up.

|  | Total (n = 96) | Worse (n = 15 ) | Stable (n = 56) | Improved (n = 25) | ANOVA (p) |
| --- | --- | --- | --- | --- | --- |
| **Sub-components** |  |  |  |  |  |
| Total fistula length, median, mm (IQR) | 47.0 (30.0 – 80.0) | 59.0 (22.0 - 110) | 47.0 (31.0 – 85.0) | 45 (32.0-77.5) | 0.75 |
| Hyperintensity on T1, present, n (%) | 80 (83.3%) | 10 (66.7%) | 47 (83.9%) | 23 (92.0%) | 0.15 |
| Hyperintensity on T2, present, n (%) | 87 (90.6%) | 13 (86.7%) | 51 (91.1%) | 23 (92.0%) | **0.04** |
| Rectal wall involvement, present |  |  |  |  | 0.48 |
| Normal, n (%) | 62 (64.6%) | 7 (46.7%) | 39 (69.6%) | 16 (64.0%) |  |
| Thickened, n (%) | 17 (17.7%) | 5 (33.3%) | 9 (16.1%) | 3 (12.0%) |  |
| Increased signal intensity, n (%) | 17 (17.7%) | 3 (20.0%) | 8 (14.3%) | 6 (24.0%) |  |
| Inflammatory mass, present, n (%) | 59 (61.5%) | 11 (73.3%) | 35 (62.5%) | 13 (52.0%) | 0.62 |
| Genitourinary tract extension |  |  |  |  | 0.051 |
| Anovaginal tract, n (%) | 13 (13.5%) | 3 (20.0%) | 9 (16.1%) | 1 (4.0%) |  |
| Peno-scrotal-perineum tract, n (%) | 8 (8.3%) | 0 (0%) | 3 (5.4%) | 5 (20.0%) |  |
| **MRI Index Scores\*** |  |  |  |  |  |
| MAGNIFI-CD, median, total score (IQR) | 14 (10 – 18.3) | 15.5 (7.25 – 19.0) | 14 (11.0 – 19.0) | 14 (7.0 - 16.0) | 0.81 |
| mVAI, median, total score (IQR) | 8.3 (4.7 - 12.3) | 9.7 (3.5 - 12.9) | 8.0 (5.0 - 13.0) | 7.9 (4.5 – 10.3) | 0.59 |
| PEMPAC, median, total score(IQR) | 26 (15 – 31.2) | 31 (11 - 33) | 26 (15.0 – 31.0) | 13 (13.0 - 24.0) | 0.58 |
| **MRI Index Score Changes<sup>†</sup>** |  |  |  |  |  |
| ΔMAGNIFI-CD, mean (σ) | -0.7 (6.4) | 1.0 (8.5) | -0.4 (6.0) | -2.3 (5.9) | **0.04** |
| ΔmVAI, mean (σ) | -1.2 (4.8) | -0.4 (6.1) | -0.9 (4.5) | -2.4 (4.5) | **0.03** |
| ΔPEMPAC, mean (σ) | -1.5 (8.9) | 0.0 (9.9) | -0.8 (8.1) | -4.1 (9.8) | **0.03** |
| ΔAveraged score, mean (σ)<sup>#</sup> | -0.0473 (0.220) | -0.00354 (0.273) | -0.0340 (0.205) | -0.108 (0.217) | **0.023** |

Abbreviations: Interquartile range (IQR), magnetic resonance novel index for fistula imaging in CD (MAGNIFI-CD), modified Van Assche Index (mVAI), pediatric MRI-based perianal Crohn's disease (PEMPAC)
\*Baseline MRI pelvis characteristics
<sup>†</sup>Delta calculated by 1st follow-up MRI pelvis score minus baseline MRI pelvis score
<sup>#</sup>Delta averaged MRI score defined as ΔMAGNIFI-CD/25 + ΔmVAI/19.5 + ΔPEMPAC/41

In a linear mixed effect model, only male sex (−0.31, 95% CI −0.60 to −0.02) was associated with improvement in Geldof classification (**Table 3**). When applied to the linear mixed model, neither observed changes in composite MRI scores nor MRI subcomponents identified by ANOVA showed any significant effect. Using change of averaged MRI score achieves slightly better prediction than using the three MRI subcomponents scores. The final model explained only a small fraction of the variance in change in Geldof classification in the cohort overt time (R^2^ = 0.10).

**Table 3:** Linear mixed effect model evaluating variables effecting change in Geldof Classification.

|  | Geldof<br>Change<sup>*</sup> | 95% CI | p |
| --- | --- | --- | --- |
| Male sex | -0.311 | (-0.603, -0.019) | 0.037 |
| History of perianal surgery | -0.194 | (-0.51, 0.122) | 0.229 |
| Hyperintensity on T2 (MRI) | 0.271 | (-0.195, 0.737) | 0.256 |
| Upper GI Involvement | 0.158 | (-0.193, 0.509) | 0.376 |
| ΔAveraged MRI score<sup>#</sup> | 0.257 | (-0.494, 1.008) | 0.502 |
| R<sup>2</sup>=0.10 |  |  |  |

<sup>\*</sup>Effect estimate of change in Geldof Classification, positive change indicating worsening class (1 < 2a < 2b < 2c-i = 2c-ii < 3 < 4a = 4b)
<sup>#</sup>Delta averaged MRI score defined

## Discussion

We present a real-world application of the Geldof classification system to patients with CD using a serial pMRI cohort. Our findings highlight the chronic relapsing and remitting nature of CD-PAF and the dynamic flexibility of the Geldof classification system which incorporates synchronization of patient and clinician goals in decision-making with a combined medical and surgical approach^14^. This is most evident by the observation that approximately half of patients (52%) changed their Geldof classifications at least once during the median 1.8 years study period.

Incidental and minimally symptomatic disease (Class 1) was observed in a minority (8.3%), consistent with other studies that have reported approximately 12% having incidental CD-PAF^19^. Chronic symptomatic fistula (Class 2), and specifically the cycle between Class 2b (symptom control) and Class 2a (repair) dominate the cohort. At initial enrollment, very few patients (5.2%) were amenable to combined medical and surgical therapy aimed at fistula closure. The transition from Class 2b to 2a was the most frequently observed and involved 1 in 5 patients whose initial therapeutic target was symptom control. These data help to further discussions regarding therapeutic goals and how these preferred transitions can be achieved through both proactive and synchronized medical-surgical approaches in multidisciplinary settings and through innovative clinical trials designed for CD-PAF.

The Geldof classification system also achieves the authors’ stated goal of emphasizing the risk of disease progression. This was most evident by the nearly one quarter of patients who had severe uncontrolled symptoms recommended for diverting ostomy or proctectomy (Classes 2c – 4) during the relatively short study period with only a select few (4/18) who later underwent ostomy takedown. Prior historical cohorts have described progression to permanent diverting ostomy between 30-50% of patients with perianal CD over longer periods of observation^20, 21^. A recent study evaluating diverting ostomy in the post-biologic era found that in a cohort of 68 diversions without proctectomy, 31% underwent ostomy take down while 22% ultimately required proctectomy^22^.

Our ability to predict transitions in the classes was limited. Between baseline and follow-up pMRI, there was significant variance in male sex, upper gastrointestinal involvement, prior perianal surgeries, and hyperintensity on T2 imaging when comparing patients whose Geldof classification improved, remained stabled, or worsened. On linear mixed effect modeling, male sex was significantly associated with improvements in the Geldof classification. The distribution of male and female patients is variable between different previously reported surgical corhorts^23–25^. The lack of consistent objective criteria for surgical intervention and the role of patient preference in Geldof classification may suggest sources of underlying bias between repair and symptom control groups, however, further study is required to validate or refute our findings. In our cohort, there was a numerically higher proportion of ano-vaginal fistula in females in the worsened group (3/9, 33.3%) compared to stable (9/30, 30.0%) and improved (1/7, 14.8%) however the association was not statistically significant (p = 0.051) in univariate analysis and was not included in the final linear mixed model.

There was no significant variation in baseline MRI index scores including MAGNIFI-CD, mVAI, or PEMPAC between those who improved and those that did not. There was, however, significant variation in the change in each index between the first and second MRI, with those who improved having the highest decreases in total score. Studies evaluating these scores to predict responsiveness to therapy and long-term outcomes are ongoing^26^.

Our study had multiple limitations. Assignment of Geldof classification requires input of a multidisciplinary care team including gastroenterologists, colorectal surgeons, and radiologists as well as patient-centered input regarding goals of care. Retrospective assignments at discrete cross-sectional intervals may result in a source of bias. We attempted to mitigate by including all documented clinician and patient input within a 6- month interval around baseline and follow-up pMRI dates and objective scoring of disease activity on pMRI using validated scoring systems by fellowship-trained abdominal radiologists. We further attempted to mitigate this through a defined protocol for assignment of classes and training for the assignor with expert feedback prior to final assignment of classes. The proposed system relies upon subjective clinician impression of disease severity and inter-rater reliability has not yet been evaluated. We also chose not to evaluate outcomes associated with different Geldof classifications as classes are associated with expected surgical and clinical outcomes by definition and represent a source of significant confounding. In addition, in its proposed form, there are no descriptors to encapsulate previously treated patients with improved/resolved symptomatic disease with or without radiographic resolution. Additional clinical and radiographic measures are required for description of treatment response and long-term closure.

In conclusion, our study highlights the Geldof consensus classification’s ability to reflect the chronic and progressive nature of perianal fistulizing CD beyond the previous static clinical characterizations. However, further prospective study and data generation at other IBD centers is required for external validation and generalizability. Variables that best predict changes in this scheme remain unknown and maybe best defined in a prospective study design. Available MRI indices are limited in their ability to predict future changes in classification, however improvement in scores over time was associated with transition to lower Geldof classification.

## Supporting information

Supplementary Appendix for review

## Data Availability

Data available upon reasonable request

## Statement of Data, Analytic Methods, and Study Materials

Data, analytic methods, and study materials will be made available upon reasonable request.

## Authors’ Contributions

Matthew Schroeder: Study design, Data extraction, interpreting the data, and initial drafting manuscript.

Suha Abushamma: Data extraction and critical review of the manuscript.

Alvin T. George: Data extraction and critical review of the manuscript.

Ravella Balakrishna: Data extraction and critical review of the manuscript.

John Hickman: Data extraction and critical review of the manuscript.

Anusha Elumalai: Interpreting the data, and critical review of the manuscript.

Paul Wise: Interpreting the data, and critical review of the manuscript.

Maria Zulfiqar: Interpreting the data, and critical review of the manuscript.

Daniel R Ludwig: Interpreting the data, and critical review of the manuscript.

Anup Shetty: Interpreting the data, and critical review of the manuscript.

Satish E. Viswanath: Interpreting the data, and critical review of the manuscript.

Chongliang Luo: Data analysis, Interpreting the data, and critical review of the manuscript.

Shaji Sebastian: Interpreting the data, and critical review of the manuscript.

David H Ballard: Study concept, design and supervision, interpreting the data, and initial drafting of the manuscript.

Parakkal Deepak: Study concept, design and supervision, interpreting the data, and initial drafting of the manuscript.

