## Supplementary Appendix for review for "Geldof Expert Consensus Classification of Perianal Fistulizing Crohn’s Disease: A Real-World Application in a Serial Fistula MRI Cohort"

Appendix A: Assignment of Geldof Classifications

Class 1: Minimal disease

- Geldof et al. “Patients with minimal symptoms and anorectal disease burden, requiring minimal intervention over time”
    - No prior history of surgical correction of perianal fistulae
    - No recommended surgical correction of perianal fistulae
    - Incidentally detected perianal fistulae on cross sectional imaging without symptoms OR
    - No prior symptomatology beyond (“patients perceive their symptoms to not be bothersome [and do] not want intervention”)
        - No or minimal mucous discharge
        - No activity restriction
        - No or mild discomfort
        - No restriction of sexual activities
        - No or minimal perianal induration
        - Less than three perianal fistulae
    - No prior history of higher Geldof et al classification

Class 2: Chronic symptomatic fistulae

- Class 2a: Repair
    - Geldof et al. “Symptomatic fistulae suitable for combined medical and surgical closure or repair (including seton removal) and patient goal is fistula closure”
        - Symptomatic fistulae (at least moderate mucous or purulent discharge, activity restriction, moderate discomfort, restriction of sexual activity, moderate induration, or greater than three perianal fistulae)
        - Ongoing or anticipated medical therapy (antibiotics, immunosuppressants, biologic, MSC) and surgical/procedural therapy (fistulotomy, ligation of fistulotomy tracks, fibrin glue, bioprosthetic plug) directed at closure of fistula
- Class 2b: Symptom control
    - Geldof et al. “Chronic symptoms related to fistulae (pain and discharge) that affect quality of life. Fistulae are currently unsuitable for surgical repair, and patient goal is symptom control”
        - Symptomatic fistulae at affect quality of life (at least moderate mucous or purulent discharge, activity restriction, moderate discomfort, restriction of sexual activity, moderate induration, or greater than three perianal fistulae)
        - Ongoing or anticipated medical therapy (antibiotics, immunosuppressants, biologic) and surgical therapy directed at improving patient symptoms who are not candidates for surgical repair
        - Therapy may be direct towards improving condition and anatomy so that surgical correction may be considered in the future (2b -> 2a)
- Class 2c-i: Early and rapidly progressive disease
    - Geldof et al. “early and rapidly progressive disease destructive to the perineum or to quality of life (or both), such that early intervention with defunctioning ostomy and sometimes early proctectomy is required”

        - Development and progression of symptom within 12 months of symptom onset
        - Severe symptomatic fistulae with at least (substantial discharge, marked discomfort, marked limitations, or marked induration) impairing quality of life
        - Disease progression due to primary or secondary non-response despite adherence to medical therapy (antibiotics, immunosuppressants, biologic, MSC) and surgical intervention for at least 3-6 months
        - Defunctioning ostomy or early proctectomy recommended by provider

  Class 2c-ii: Gradually debilitating disease

    - Geldof et al. "gradually debilitating symptomatic fistulae unsuitable for surgical repair, which cause severe symptoms, limiting quality of life so markedly that defunctioning ostomy is required to restore quality of life. Patient goal is symptom control"
        - Disease persistence and progression for at least 12 months
        - Severe symptomatic fistulae (at least substantial discharge, marked discomfort, marked limitations, or marked induration) impairing quality of life
        - Defunctioning ostomy is recommended by provider

<u>Class 3: Severe disease with exhausted perinium and adverse feature</u>

- Geldof et al. "Patients who have severely symptomatic disease (despite defunctioning), with irreversible perineal destruction, or symptoms limiting quality of life so markedly that proctectomy is required"

  "Patients who have severely symptomatic disease (despite defunctioning), with irreversible perineal destruction, or symptoms limiting quality of life so markedly that proctectomy is required"

    - Persistent severe symptomatic fistulae (at least substantial discharge, marked discomfort, marked limitations, or marked induration)
    - Proctectomy recommended by provider

<u>Class 4: Perineal symptoms after proctectomy</u>

- Class 4a: Repair
    - Geldof et al. "Patients with symptomatic sinuses or wounds suitable for combined medical and surgical closure or repair and whose goal is sinus closure"
        - History of proctectomy
        - Recurrent symptoms with either symptomatic sinuses or wounds
        - Ongoing or anticipated optimized medical therapy in combination with surgical correction targeted at repair of chronic sinus.
- Class 4b: Symptom Control
    - Geldof et al. "Patients with chronic symptoms related to their sinuses or wounds that affect their quality of life, whose sinuses or wounds are unsuitable for surgical repair, or whose goal is symptom control"
        - History of proctectomy
        - Recurrent symptoms with either symptomatic sinuses or wounds
        - Ongoing or anticipated optimized medical therapy in combination with surgical correction targeted at repair of chronic sinus who are not candidates for surgical repair.

Appendix B: Score Assignments of Previously Established MRI Indices

| | mVAI [2017] | | MAGNIFI-CD [2019] | | PEMPAC [2022] | |
|---|---|---|---|---|---|---|
| **Sub-sores:** | Item Score | | Item Score | | Item Score | |
| **Number of fistulas** | | | None | 0 | None | 0 |
| | | | Single, unbranched | 3 | Single | 4 |
| | | | Complex | 6 | Multiple | 8 |
| **Location** | | | | | None | 0 |
| | | | | | Intersphincteric | 3 |
| | | | | | Transsphincteric | 6 |
| | | | | | Extrasphincteric | 9 |
| | | | | | Transsphincteric/ intersphincteric | 12 |
| **Extension** | Absent | 0 | Absent | 0 | | |
| | Infralavator | 1.5 | Horseshoe | 2 | | |
| | Horseshoe | 3 | Intralavator or supralevator | 4 | | |
| | Supralavator | 4.5 | | | | |
| **Length of fistulas** | | | < 2.5 cm | 0 | None | 0 |
| | | | 2.5 - 5 cm | 2 | 0.1 - 2.5 cm | 2 |
| | | | > 5 cm | 4 | 2.6 - 5.0 cm | 4 |
| | | | | | > 5.0 cm | 6 |
| **Dominant feature** | Fibrous | 0 | Fibrous | 0 | | |
| | Granulation tissue | 1.2 | Granulation tissue | 2 | | |
| | Fluid or pus | 2.4 | Fluid or pus | 4 | | |
| **Rectal wall involvement** | Normal | 0 | | | | |
| | Thickened | 1 | | | | |
| | Increased signal intensity | 2 | | | | |
| **Inflammatory mass** | Absent | 0 | Absent | 0 | Absent | 0 |
| | Diffuse | 1.2 | Focal | 1 | Present | 11 |

| | | | | | | |
|---|---|---|---|---|---|---|
| | Focal | 2.4 | Diffuse | 2 | | |
| | Small collection | 3.6 | Small collection | 3 | | |
| | Medium collection | 4.8 | Medium collection | 4 | | |
| | Large collection | 6 | Large collection | 5 | | |
| **Maximal T2 hyper-intensity** | Absent | 0 | | | Absent | 0 |
| | Mild | 2.3 | | | Mild | 2 |
| | Pronounced | 4.6 | | | Pronounced | 4 |
| **Hyperintensity of primary tract on postcontrast T1-weighted images** | | | Absent or mild | 0 | | |
| | | | Pronounced | 2 | | |
| **Score Range** | | **0-19.5** | | **0-25** | | **0-41** |
